# Neutralizing antibody responses to SARS-CoV-2 in a COVID-19 recovered patient cohort and their implications

**DOI:** 10.1101/2020.03.30.20047365

**Authors:** Fan Wu, Aojie Wang, Mei Liu, Qimin Wang, Jun Chen, Shuai Xia, Yun Ling, Yuling Zhang, Jingna Xun, Lu Lu, Shibo Jiang, Hongzhou Lu, Yumei Wen, Jinghe Huang

## Abstract

The COVID-19 pandemic caused by SARS-CoV-2 coronavirus threatens global public health. Currently, neutralizing antibodies (NAbs) versus this virus are expected to correlate with recovery and protection of this disease. However, the characteristics of these antibodies have not been well studied in association with the clinical manifestations in patients.

**Methods:** Plasma collected from 175 COVID-19 recovered patients with mild symptoms were screened using a safe and sensitive pseudotyped-lentiviral-vector-based neutralization assay. Spike-binding antibody in plasma were determined by ELISA using RBD, S1, and S2 proteins of SARS-CoV-2. The levels and the time course of SARS-CoV-2-specific NAbs and the spike-binding antibodies were monitored at the same time.

**Findings:** SARS-CoV-2 NAbs were unable to cross-reactive with SARS-CoV virus. SARS-CoV-2-specific NAbs were detected in patients from day 10-15 after the onset of the disease and remained thereafter. The titers of NAb among these patients correlated with the spike-binding antibodies targeting S1, RBD, and S2 regions. The titers of NAbs were variable in different patients. Elderly and middle-age patients had significantly higher plasma NAb titers (P<0.0001) and spike-binding antibodies (P=0.0003) than young patients. Notably, among these patients, there were ten patients whose NAb titers were under the detectable level of our assay (ID50: < 40); while in contrast, two patients, showed very high titers of NAb, with ID50 :15989 and 21567 respectively. The NAb titers were positive correlated with plasma CRP levels but negative correlated with the lymphocyte counts of patients at the time of admission, indicating an association between humoral response and cellular immune response.

**Interpretation:** The variations of SARS-CoV-2 specific NAbs in recovered COVID-19 patients may raise the concern about the role of NAbs on disease progression. The correlation of NAb titers with age, lymphocyte counts, and blood CRP levels suggested that the interplay between virus and host immune response in coronavirus infections should be further explored for the development of effective vaccine against SARS-CoV-2 virus. Furthermore, titration of NAb is helpful prior to the use of convalescent plasma for prevention or treatment.

**Funding:** Ministry of Science and Technology of China, National Natural Science Foundation of China, Shanghai Municipal Health Commission, and Chinese Academy of Medical Sciences

## Introduction

The outbreak of coronavirus disease 2019 (COVID-19) in December 2019 has spread around the world and become a global pandemic.^1^ The etiological agent of COVID-19 was identified as a SARS-related coronavirus designated as SARS-COV-2 coronavirus.^2,3^ As of March 27, 2020, it had caused a total of 509,164 cases of infection and resulted in 23,335 deaths worldwide.^1^ About 81% of infected patients showed only mild symptoms, but 14% of them had severe symptoms such as dyspnea, high respiratory frequency and low blood oxygen saturation. Another 5% of patients, especially those over 60, or with comorbidities, progressed to critical condition. About 3.4% of patients died from respiratory failure or multiple organ failure.^4^ Although the estimated mortality rate of COVID-19 was lower than SARS and MERS, the number of deaths associated with COVID-19 has already surpassed those of SARS and MERS owing to the extremely high transmissibility of SARS-CoV-2 coronavirus. Currently, no licensed vaccine or drugs are available to prevent or treat COVID-19 infection, and most infected patients have been treated with supportive care.

Neutralizing antibodies (NAbs) play important roles in virus clearance and have been considered as a key immune product for protection or treatment against viral diseases. Virus-specific NAbs, induced through either infection or vaccination, have the ability to block viral infection. The level of NAbs has been used as a gold standard to evaluate the efficacy of vaccines against smallpox, polio and influenza viruses.^5^ Passive antibody therapy, such as plasma fusion, was successfully used to treat infectious viral diseases, including SARS-CoV virus,^6^ influenza viruses,^7^ and Ebola virus.^8^ The efficacy of passive antibody therapy was associated with the concentration of NAbs in plasma or antibodies of recovered donors.^8^ As the global pandemic of COVID-19 proceeds, transfusion of convalescent plasma or serum from recovered patients was also considered as a promising therapy for prophylaxis of infection or treatment of disease.^9^ However, the levels and roles of SARS-CoV-2-specific NAbs in patients with COVID-19 have not been reported.

Here, we used a pseudotyped-lentiviral-vector-based neutralization assay to measure SARS-Cov-2-specific NAbs in plasma from recovered COVID-19 patients with mild symptoms. The pseudovirus (PsV) neutralization assay is a sensitive and reproducible assay. It does not produce any highly pathogenic virus, and it can be safely handled in a biosafety level 2 facility. Herein, we aimed to explore the clinical characteristics associated with the level of NAbs in recovered patients, the outcome of which may provide useful information for the development of vaccines and passive antibody therapy for the prevention and treatment of SARS-CoV-2.

## Methods

### Study design and participants

The study included a cohort of 175 adult COVID-19 patients admitted to Shanghai Public Health Clinical Center. The study was conducted under a clinical protocol approved by the Investigational Review Board in the Shanghai Public Health Clinical Center (Study number: YJ-2020-S021-01). All participants signed an informed consent approved by the IRB. All patients were diagnosed with laboratory-confirmed COVID-19 and discharged after meeting effective national treatment standards. Clinical information, including complete blood counts, blood biochemistry was collected at the time of admission.

## Materials

293T cells expressing human angiotensin converting enzyme II (ACE2) (293 T/ACE2) were obtained from the American Type Culture Collection (ATCC; Manassas, VA, USA) and were cultured in Dulbecco’s modified Eagle’s medium (DMEM) with 10% fetal bovine serum (FBS). The three domains of SARS-CoV-2 spike (S) protein, including S1 and S2 subunits, as well as RBD, were purchased from Sino Biological Company (Beijing, China). The expression plasmids for SARS S protein pcDNA3.1-SARS-S (ABD72979.1) and SARS-CoV-2 S protein pcDNA3.1-SARS-CoV-2-S (NC_045512) were synthesized by Genscript. The VSV-G envelope eukaryotic expression vector pHEF-VSVG and the HIV-1 Env-deficient luciferase reporter vector pNL4-3. Luc. R-E- were obtained through the NIH AIDS Reagent Program.

### Neutralization assay

Neutralization activity of plasma from COVID-19 patients was measured using a single-round PsV infection of 293 T/ACE2 cells. PsVs of SARS-CoV-2, SARS-CoV and VSV-G virus were generated by co-transfection of 293T cells with pNL4-3.Luc.R-E-backbone and viral envelope protein expression plasmids pcDNA3.1-SARS-CoV-2-S, pcDNA3.1-SARS-S or pHEF-VSVG. PsVs could infect the same cells as those infected by SARS-CoV-2 or SARS-CoV viruses.^10,11^ The neutralization assay was performed in accordance with the following steps. First, 293 T/ACE2 cells were seeded in a 96-well plate at a concentration of 10^4^ cells per well and cultured for 12 hours. Then, ten μl heat-inactivated plasma were five-fold serially diluted with DMEM with 10% FBS and mixed with 40 μl of PsV. The mixture was added to cultured 293 T/ACE2 for infection. The culture medium was refreshed after 12 hours and incubated for an additional 48 hours. Assays were developed with a luciferase assay system (Promega), and the relative light units (RLU) were read on a luminometer (Perkin Elmer). The titers of NAbs were calculated as 50% inhibitory dose (ID50), expressed as the highest dilution of plasma which resulted in a 50% reduction of luciferase luminescence compared with virus control.

### ELISA

SARS-CoV-2 RBD, S1, or S2 protein and SARS-CoV RBD or S1 protein (1 μg/ml) was coated on a MaxiSorp Nunc-immuno 96-well plate overnight at 4 °C. Wells were blocked with 5% nonfat milk in PBS for 1 hour at room temperature, followed by incubation with 1:400 diluted sera or serially diluted sera in disruption buffer (PBS, 5% FBS, 2% BSA, and 1% Tween-20) for 1 hour at room temperature. A 1:2500 dilution of horseradish peroxidase (HRP)-conjugated goat anti-human IgG antibody was added for 1 hour at room temperature. Wells were washed five times between each step with 0.2% Tween-20 in PBS. Wells were developed using ABST (Thermo) and read at 405 nm.

### Statistical analysis

Statistical analyses were carried out using GraphPad Prism 7.0. Data are indicated as medians. Differences between nominal data were tested for statistical significance by use of paired or unpaired *t* test. Correlations were calculated using standard Pearson correlation.

## Results

### Clinical Characteristics

A total of 175 COVID-19 patients had recovered and were discharged from the Shanghai Public Health Clinical Center as of February 26, 2020. Their symptoms were common or mild, and none of them was admitted to the ICU. The median age of the patients was 50 years (ranging from 16 to 85 years); 53 % of the patients were female. The median length of hospital stay was 16 days (ranging from 7 to 30 days), and the median disease duration was 21 days (9 to 34 days).

### Convalescent plasma from COVID-19 patients specifically inhibited SARS-CoV-2, but not SARS-CoV infection

We collected five plasma samples from COVID-19 patients at the time of discharge and measured their neutralizing titers against SARS-CoV-2 infection of 293T/ACE2 cells. All five plasma showed a concentration-dependent inhibition of SARS-CoV-2 PsV infection of 293T/ACE2 cells (Figure 1A). Plasma with high titers of NAbs showed higher titers of SARS-CoV-2 RBD, S1, and S2-specific binding antibodies (Figure 1B). Moreover, plasma from these patients also showed cross-binding to SRAS-CoV RBD and S1 regions (Figure 1C), but the binding to SARS-CoV S protein was not consistent with that to SARS-CoV-2 S protein. Furthermore, plasma from COVID-19 patients could not inhibit SARS-CoV infection in PsV neutralization assay. 26 plasma samples from COVID-19 patients, which showed strong SARS-CoV-2 neutralizing activities (Figure 1D), could neither neutralize SARS-CoV PsV infection nor the control VSV-G PsV infection (Figure 1E). These results suggest that SARS-CoV-2 was able to stimulate SARS-CoV cross-binding antibodies. However, it was unable to induce the cross-neutralizing antibodies against SARS-CoV. These results suggested that the epitope or immunogenicity between SARS-CoV-2 and SARS-CoV were different.

**Figure 1.**
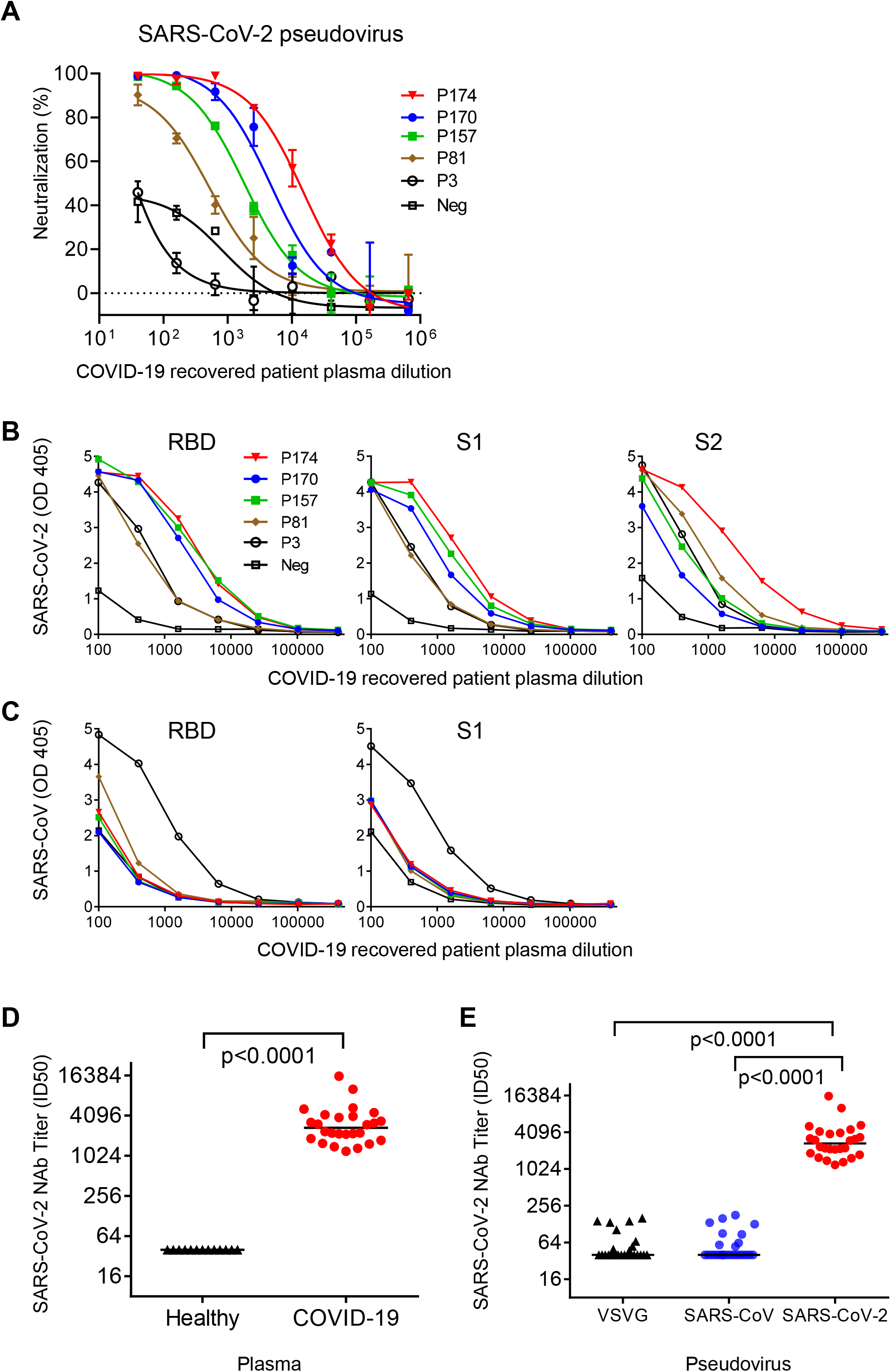
Plasma from COVID-19 recovered patients specifically inhibited SARS-CoV-2 infection but not SARS-CoV virus. (A) Plasma from five COVID-19 recovered patients inhibited infection of SARS-CoV-2. Plasma from a healthy donor was used as a negative control. The assay was performed in duplicate and the median percentage of neutralization is shown. (B) Biding of COVID-19 recovered patient plasma to SARS-CoV-2 RBD, S1, and S2 proteins. (C) Biding of COVID-19 recovered patient plasma to SARS-CoV RBD and S1 proteins. (D) The SARS-CoV-2 NAbs titers of 26 plasma from COVID-19 recovered patients were compared with 13 plasma from healthy donors. P value was calculated using *t* test. (E)The titers of NAbs against VSV, SARS-CoV, and SARS-CoV-2 PsV in 26 COVID-19 recovered patient plasma were compared. P values were calculated using *t* test.

### COVID-19 patients generated SARS-CoV-2-specific NAbs and spike-binding antibodies concurrently from day 10 to 15 after infection

We monitored the kinetics of SARS-CoV-2-specific NAb development during the course of disease. The titers of NAbs were evaluated in plasma collected from six patients at different time points after the disease onset. The kinetics of NAbs development were similar among patients. The titers of NAbs in all patients were low (ID50: < 200) before day 10 post-disease onset and then increased at day 10 to 15 post-disease onset, remaining stable thereafter (Figure 2A). We also measured the binding antibodies to the different domains (RBD, S1, and S2) of SARS-CoV-2 spike protein in the plasma of these six patients. The kinetics of NAbs (right Y axis) and binding antibodies targeting RBD, S1, and S2 domains (left Y axis) were aligned with individual patients (Figure 2B). We evaluated the SARS-CoV-2-specific NAbs titers and the spike-binding antibody levels in the plasma of 175 recovered patients on the day of discharge. We observed that SARS-CoV-2-specific NAbs titers moderately correlated with spike-binding antibodies targeting RBD (r=0.51, p<0.0001), S1 (r=0.42, p<0.0001), and S2 (r=0.435, p<0.0001) (Figure 2C). These results suggested that humoral immune responses of COVID-19 patients against SARS-CoV-2 occurred on day 10 to 15 after infection. Besides RBD region, S2 domain might be the target of SARS-CoV-2-NAbs. Since binding antibodies may also play a role in viral clearance through antibody-dependent phagocytosis or antibody-dependent cellular cytotoxicity, the effect of NAbs and binding antibodies on disease progression is worth comprehensive evaluation in further study.

**Figure 2.**
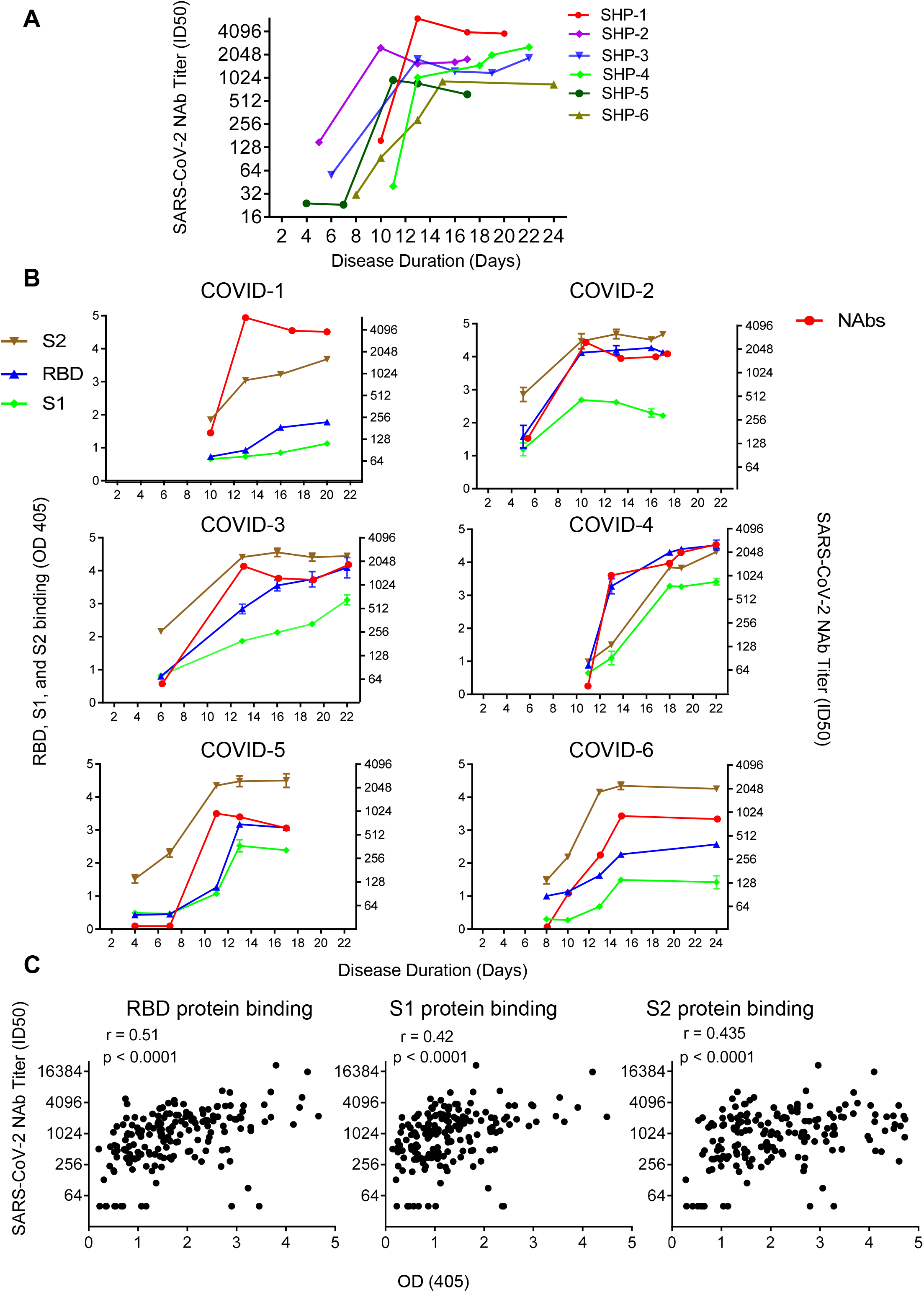
SARS-CoV-2-specific NAbs and spike-binding antibodies emerged concurrently on day 10-15 during the COVID-19 disease progression and shown correlation. (A) Kinetics of SARS-CoV-2 NAbs titers in six COVID-19 patients are shown. Plasma were collected at different time points post syndrome onset. (B) Kinetics of spike binding antibodies (left Y axis), targeting RBD (blue), S1 (green), and S2 (brown), in six COVID-19 patient plasma are shown and compared with the kinetics of Nabs titers (right Y axis, red) in the same patient. (C) The correlations between the SARS-CoV-2 NAbs titers and RBD, S1, or S2 binding antibodies levels of patients were analyzed by Pearson correlation test. 1:400 diluted plasma was incubate with RBD, S1, or S2 protein.

### About 30% of recovered patients generated very low titers of SARS-CoV-2-specific NAbs

We observed that NAb titers were variable in the plasma of 175 recovered patients. ID50s ranged from below detection limit (<40) to 21567 (Figure 3A). About 30% of recovered patients generated a very low level of NAb titers (ID50: < 500) (Figure 3A, 3B, and Supplementary Table 1), and NAb titers in ten of them were below the limit of detection (ID50: <40), though all of them were lab confirmed infected with SARS-CoV-2 (Supplementary Table 2). About 17%, 39%, and 14% showed medium-low (ID50: 500-999), medium-high (ID50: 1000-2500), and high (ID50: > 2500) NAb titers, respectively (Figure 3B). We also collected and measured the levels of NAbs in plasma from 47 of the 175 patients during the follow-up examination two weeks after discharge. As shown in Figure 3C, NAb plasma titers collected at the time of follow-up examinations did not significantly differ from those collected at the time of discharge (P=0.250, paired-*t* test). Patients who did not generate NAbs at the time of discharge did not develop NAbs thereafter. These results revealed that a proportion of patients infected with SARS-CoV-2 would recover without developing high titers of virus-specific NAbs. How these patients recovered without the help of NAbs and whether they were at risk of re-infection of SARS-CoV-2 should be further explored. Titration of NAb is helpful prior to the use of convalescent plasma for prevention or treatment.

**Figure 3.**
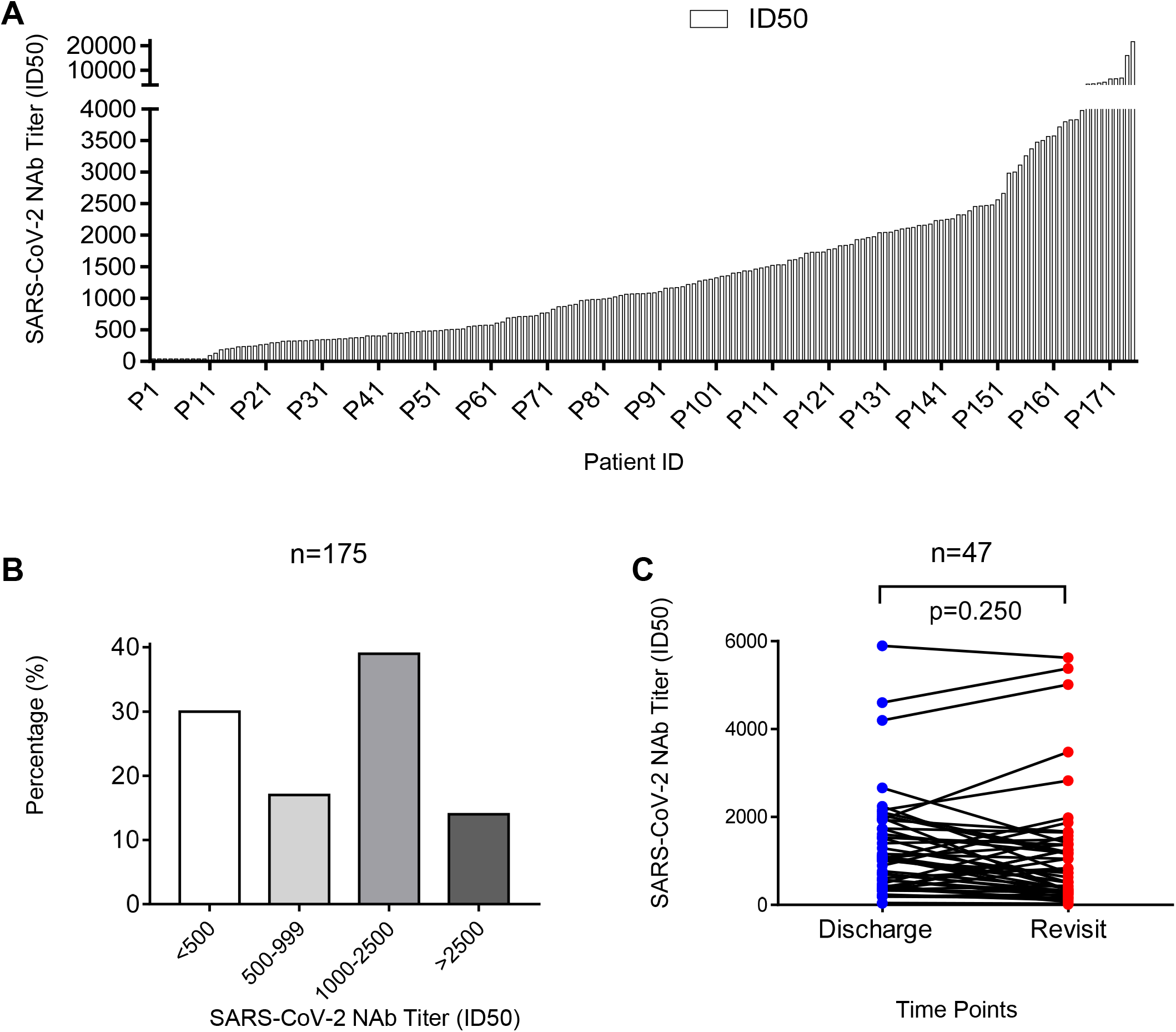
COVID-19 recovered patients developed variable levels of SARS-CoV-2 specific NAbs.. (A) SARS-CoV-2 NAb titers (ID50) of 175 COVID-19 recovered patient plasma collected on the day of discharge were measured in a PsV neutralization assay. (B) Percentages of patients with low (ID50: <500), medium-low (ID50: 500-999), medium-high (ID50: 1000-2500), and high (ID50: >2500) titers of SARS-CoV-2-specific NAbs are shown. (C) NAbs titers of 47 COVID-19 recovered patient plasma collected on the day of discharge and the subsequent visit in two weeks were compared. P value was calculated using *t* test.

### Elderly and middle-age recovered COVID-19 patients developed higher levels of SARS-CoV-2-specific NAbs

We observed that elderly patients were more likely to induce higher titers of NAbs than younger patients. As shown in Figure 4A, the patients were divided into three groups based on their age, young (15-39 years), middle-age (40-59 years) and elderly (60-85 years). Patient numbers from each group were similar (55, 64 and 56) (Supplementary Table 3). NAb titers of elderly and middle-age recovered patients were significantly higher than those of young recovered patients (p<0.0001 and p<0.0001, *t* test) (Figure 4A), and the corresponding median ID50s were 1537, 1255, and 488, respectively (Figure 4A). A moderate positive correlation was also observed between age and NAb titers (r=0.436, P<0.001, Pearson) (Figure 4C), confirming the important role of age in the generation of NAbs. Elderly and middle-age recovered patients had significantly higher levels of spike-binding antibodies, targeting RBD (p<0.0001 and p=0.0094, *t* test), S1 (p=0.0003 and p=0.0035, *t* test), and S2 (p=0.0003 and p=0.0019, *t* test) than those of young recovered patients (Figure 4C). However, no difference was observed between patients’ ages and the length of stay in hospital (Figure 4D). These results indicated that high level of NAbs might be useful to clear the viruses and helpful for the recovery of elderly and middle-age patients.

**Figure 4.**
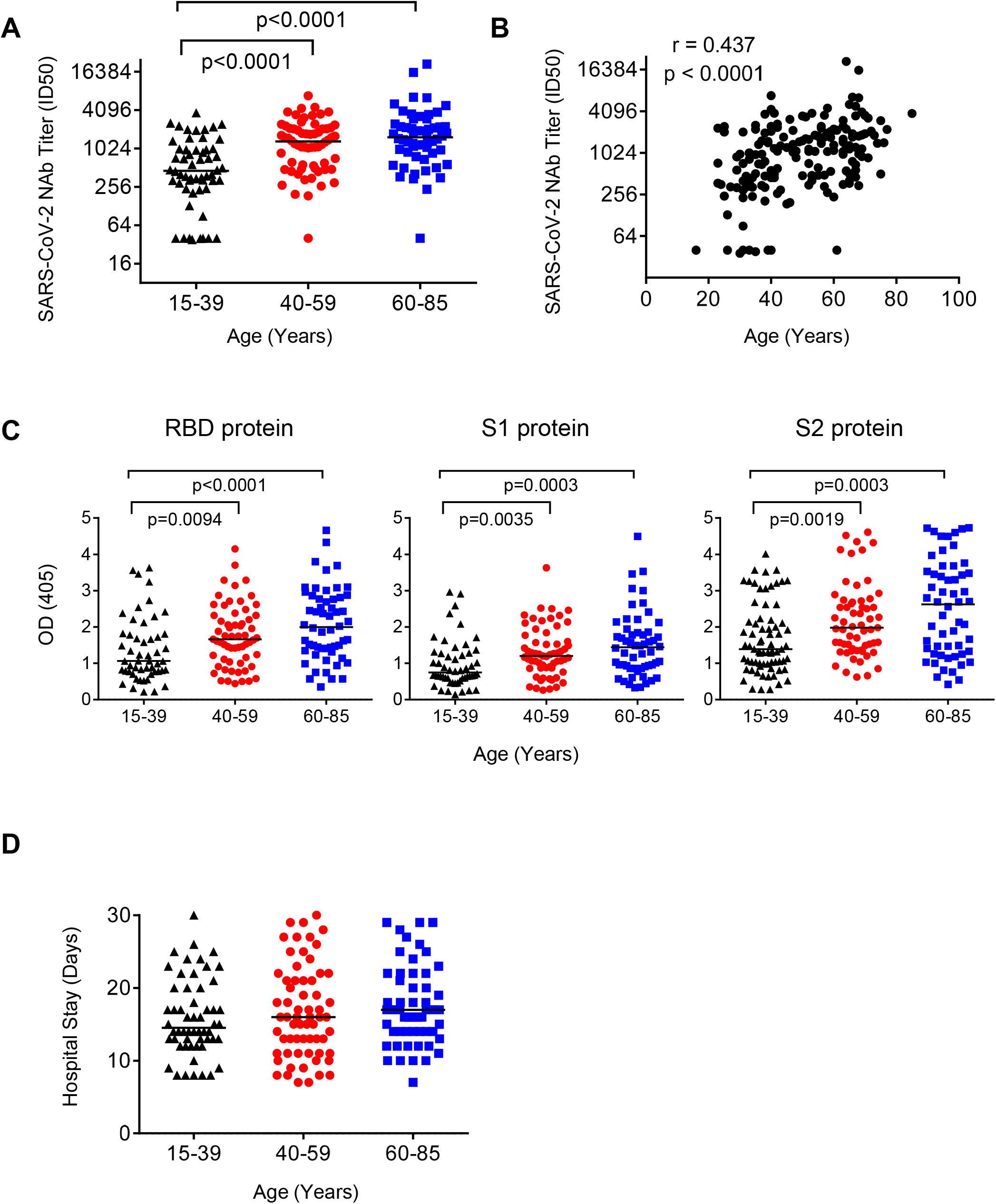
Elderly and middle-age recovered COVID-19 patients developed higher levels of SARS-CoV-2-specific NAbs than young recovered patients. (A) NAbs titers of young (15-39 years), middle-age (40-59 years), and elderly (60-85 years) patients were compared. P values were calculated using *t* test. (B) The correlation between ages of patients and the titers of SARS-CoV-2-specific NAbs was analyzed by Pearson correlation test. (C) RBD, S1, or S2 binding antibodies levels of young, middle-age, and elderly recovered COVID-19 patients were compared. P values were calculated using *t* test.

### COVID-19 recovered patients age and SARS-CoV-2-specific NAbs titers negatively correlated with lymphocyte count and positively correlated with CRP levels on admission

Older age was usually associated with poor outcome among COVID-19 patients^12^. Consistent with the previous reports, the elderly and middle-age patients in this cohort had lower lymphocyte counts (r= −0.389, p<0.0001, Figure 5A left) and higher CRP level (r= −0.432, p<0.0001, Figure 5A right) than young patients on admission (Figure 5A left and right). However, none of the patient progressed into severe conditions, and no significant difference was observed between age and length of hospital stay among these patients (Figure 4D). Interestingly, we observed that the NAb titers negatively correlated with blood lymphocyte counts (r= −0.44, p<0.0001, Figure 5B left) and positively correlated with blood CRP levels (r= 0.5, p<0.0001, Figure 5B right), suggesting that the humoral response might play an important role when cellular response was dysfunction or impaired.

**Figure 5.**
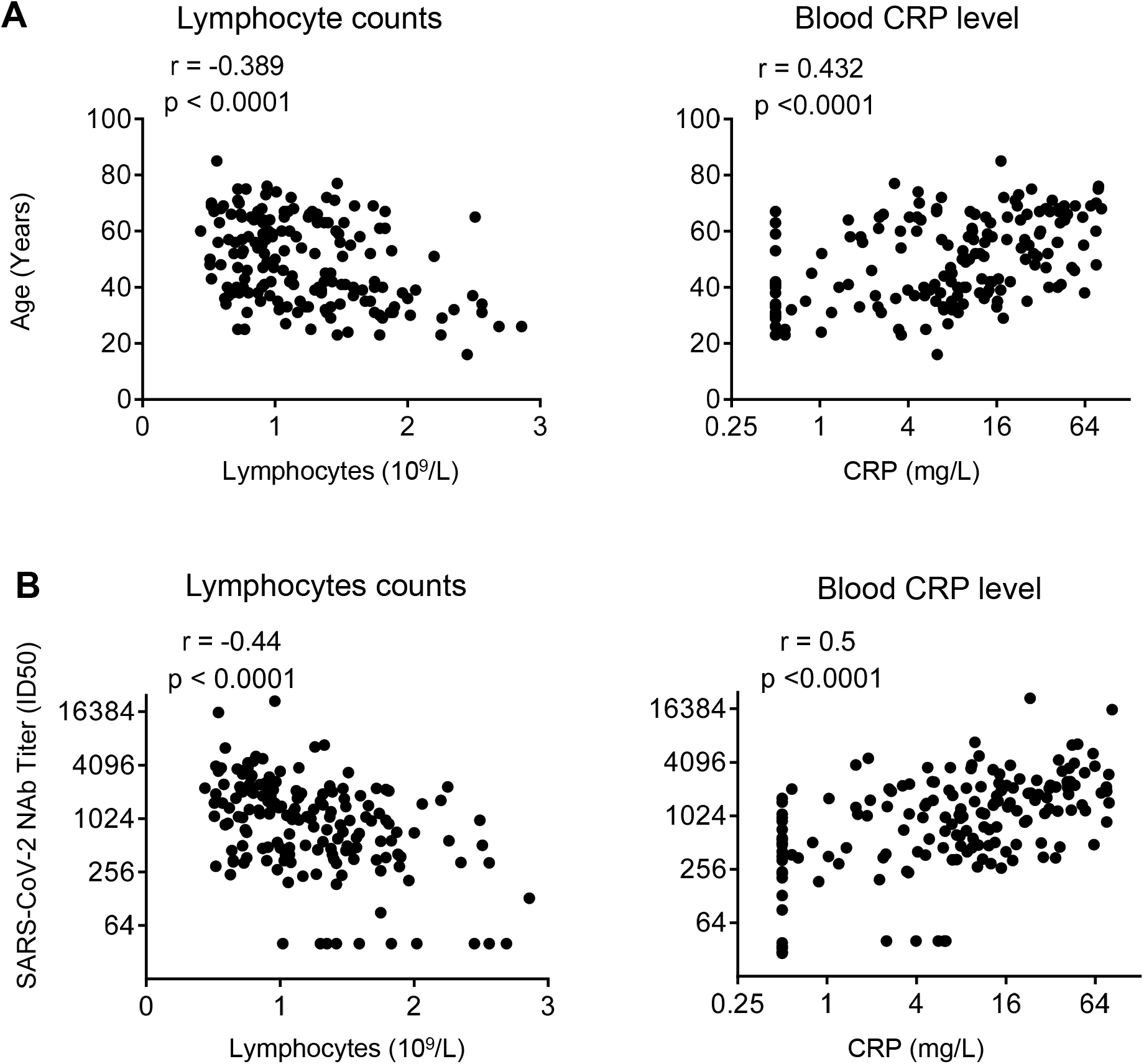
Age and SARS-CoV-2-specific NAb levels negatively correlated with lymphocyte count and positively correlated with CRP levels of patients on the time admission. (A) The correlations between patient age and lymphocyte counts (left) or C-reactive protein (CRP) level (right) on admission were analyzed by Pearson correlation test. (B) Correlations between SARS-CoV-2-specific NAb titers and lymphocyte count (left) or CRP level (right) of patients were analyzed by Pearson correlation tests. The reference range for lymphocyte counts is 1.1-3.2 X10^9^ /L and for blood CRP is less than 3mg/L.

## Discussion

Spread of the COVID-19 global pandemic highlights the urgent need to develop effective treatments or vaccines against SARS-CoV-2 infection. NAbs have been considered as an effective drug to treat or prevent virus infection. Here we evaluated the level of NAbs in recovered patients of COVID-19 by using a PsVs neutralization assay, which has been extensively used for the evaluation of NAbs for many highly pathogenic viruses, including Ebola,^13^ highly pathogenic influenza virus,^14,15^ SARS-CoV,^16^ and MERS-CoV.^17^ The PsVs neutralization assay was also used for the evaluation of NAbs for SARS-CoV-2 in some recent reports,^11,18,19^ generating consistent results compared with traditional plaque reduction neutralization assay.^18^

We found that most COVID-19 patients developed SARS-CoV-2-specific NAbs at the convalescent phase of infection. The titers of NAbs reached their peak at 10 to 15 days after disease onset and remained stable thereafter in patients. Antibodies targeting on different domains of S protein, including S1, RBD, and S2, may all contribute to the neutralization.

Conserved epitopes may exist between SARS-CoV-2 and SARS-CoV since they share 77.2% identical amino acids in their spike proteins.^2^ Few reports have demonstrated that SARS-CoV-specific monoclonal NAbs could cross-neutralize SARS-CoV-2 PsV infection,^3,11,18^ Even though plasma from COVID-19 patients showed cross-binding to SARS-Cov, they did not neutralize SARS-CoV, indicating that the antigenicity of SARS-CoV-2 is different from that of SARS-CoV. Evidence deduced from this study only suggested that cross-neutralizing antibodies targeted the conserved epitopes of SARS-CoV and SARS-CoV-2 may not be easily elicited during the infection of COVID-19, making this a potential line of advanced study.

It is also noteworthy that the levels of NAbs in patients were variable. About 30% of patients failed to develop high titers of NAbs after COVID-19 infection. However, the disease duration of these patients compared to others was similar. Notably, there were ten recovered patients whose NAb titers were very low, under the detectable level of this study (ID50: <40), suggesting that other immune responses, including T cells or cytokines, may contribute to the recovery of these patients. Whether these patients were at high risk of rebound or reinfection should be explored in further studies. On the other hand, two patients had very high titer of NAbs, which were over ID50: 15989 and 21567 respectively, but did not show any antibody-related adverse reactions.

The NAbs titers in patients were also observed to be correlated with the age of the patients. Elderly patients had significantly higher titers of NAbs than younger patients. Age has been reported as an important predictor of adverse disease outcome after infection with coronavirus, including SARS-CoV^20^, MERS-CoV^21^ and SARS-CoV2^12^. Previous studies in SARS-CoV-infected macaques revealed that aged macaques induced elevated innate immune response, resulting in more severe pathology than young adult macaques^22^. The elderly patients in this cohort also had higher blood CRP level and lower lymphocyte counts at the time of admission, indicating the induction of stronger innate immune response than younger patients. High level of NAbs may be a result of strong immune response in these elderly patients. Whether the high level of NAbs protect these patients from progression into severe and critical conditions is worthy of comprehensive evaluation. Further study of the immunological characteristics of COVID-19 patients may reveal key determinants in the generation of NAbs and effective cell-mediated immune responses, which is important for the development of an effective vaccine against SARS-CoV-2 virus.

This study is preliminary and has several limitations. First, viral RNA was not detectable in patients’ blood. Owing to the lack of respiratory specimens, information about the kinetics of viral loads was not available. Second, patients in severe and critical condition were excluded from the study because they received passive antibody treatment before sample collection. Thus, we were not able to directly evaluate the effect of NAbs on virus clearance or disease progression of COVID-19 patients in this study. A further comprehensive study should be made to address the question.

To the best of our knowledge, this is the first report about NAbs drawn from the plasma of a COVID-19 recovered patient cohort, potentially providing useful information for passive antibody therapy and vaccine development against SARS-CoV-2 virus. The highly variable levels of NAbs in the patients of COVID-19 indicated that convalescent plasma and serum from recovered donors should be titrated before use in passive antibody therapy, an easy task that can be performed using the PsV neutralization assay. Correlation of NAbs titers with the age, lymphocyte counts and blood CRP levels of patients also lays the groundwork for further study to explore the mechanism of NAbs development in COVID-19 patients.

## Data Availability

The authors confirm that the data supporting the findings of this study are available within the article and its supplementary materials.

## Contributions

JH, FW, and YW conceived and designed the experiments. JH, FW, AW, ML, QW, and YZ performed the experiments. JH, FW, SX, and LL constructed the SARS-CoV-2 PsV plasmid. FW, JH, HL, JC, YL, QW, and JX collected the samples of recovered patient and clinical information. JH, FW, YW, and SJ analyzed the data and wrote the manuscript.

## Acknowledgments

This work was supported by the National Major Science and Technology Projects of China (2017ZX10202102 and 2018ZX10301403), National Natural Science Foundation of China (31771008), Hundred Talent Program of Shanghai Municipal Health Commission (2018BR08), and Chinese Academy of Medical Sciences (2019PT350002).

## Figure legends

**Supplementary Table 1.** Clinical characteristics of COVID-19 recovered patients with low, medium-low, medium-high, and high titers of SARS-CoV-2-specific NAbs

| **Patient Information** | **Low<sup>a</sup>** | **Medium-low<sup>a</sup>** | **Medium-high<sup>a</sup>** | **High<sup>a</sup>** |
| --- | --- | --- | --- | --- |
|  | **<500** | **500-999** | **1000-2500** | **>2500** |
| Recovered Patient No. | 52 (30%) | 29 (17%) | 69 (39%) | 25 (14%) |
| Male | 19 (23%) | 13 (16%) | 36 (44%) | 14 (17%) |
| Female | 33 (35%) | 16 (17%) | 33 (35%) | 11 (12%) |
| Median Age (Years) | 38 (16-68) | 42 (23-75) | 56 (23-77) | 63 (35-85) |
| Length of Stay (Days) | 14.5 (8-29) | 15 (7-28) | 16 (8-30) | 18 (10-29) |
| Disease Duration (Days) | 20 (9-33) | 21 (16-31) | 22 (11-34) | 23 (13-32) |
| Median NAb titers (ID50) | 327 (40-488) | 715 (504-989) | 1642 (1004-2482) | 3800 (2560-21567) |

<sup>a</sup> SARS-CoV-2-specific NAb titer (ID50) values < 500 were defined as low levels, values between 500 and 999 were defined as medium-low levels, values between 1000 and 2500 were defined as medium-high levels, and values >2500 were defined as high levels.

**Supplementary Table 2.** Clinical characteristics of ten COVID-19 recovered patients with undetectable level of SARS-CoV-2 specific NAbs.

| ID | Age<br>(Years) | Gender | ID50<sup>a</sup> | ID80<sup>a</sup> | Length of<br>Hospital<br>(Days) | Disease<br>Duration<br>(Days) | Temp<br>(°C) | Viral<br>RNA<br>tests | Symptoms |
| --- | --- | --- | --- | --- | --- | --- | --- | --- | --- |
| P1 | 30 | F | <40 | <40 | 22 | 31 | 37.8 | + | fever and stuffy nose |
| P2 | 35 | F | <40 | <40 | 17 | 22 | 37.6 | + | Cough, sore muscles, and stuffy nose |
| P3 | 16 | M | <40 | <40 | 9 | 12 | 37.7 | + | Stuffy nose, runny nose, and cough |
| P4 | 39 | F | <40 | <40 | 8 | 12 | 38.1 | + | Cough |
| P5 | 40 | M | <40 | <40 | 13 | 14 | 37.9 | + | Cough and chest pain |
| P6 | 33 | F | <40 | <40 | 13 | 15 | 37.4 | + | Fatigue |
| P7 | 61 | F | <40 | <40 | 18 | 22 | 37.2 | + | Chill |
| P8 | 39 | F | <40 | <40 | 21 | 23 | 38.1 | + | Sore throat, cough, and fatigue |
| P9 | 26 | F | <40 | <40 | 8 | 9 | 38 | + | Cough |
| P10 | 31 | F | <40 | <40 | 12 | 23 | 38.4 | + | Cough and dizziness |

<sup>a</sup> ID50, ID80: < 40 represents the NAb titers were under the detectable level in neutralization assay.

**Supplementary Table 3.** Clinical characteristics and SARS-CoV-2-specific NAb titers of young, middle-age, and elderly COVID-19 recovered patients

| Patient Information | Age Distribution (Years) |  |  |
| --- | --- | --- | --- |
|  | 15-39 | 40-59 | 60-85 |
| Recovered Patient No. | 55 (31%) | 64 (37%) | 56 (32%) |
| Male | 27 (33%) | 33 (40%) | 22 (27%) |
| Female | 28 (30%) | 31 (33%) | 34 (37%) |
| Length of stay (days) | 14 (8-26) | 16 (7-30) | 17 (7-29) |
| Disease duration (days) | 21 (9-32) | 21 (11-34) | 22 (15-33) |
| Median NAb titers (ID50) | 448 (40-3717) | 1255 (40-6888) | 1537 (40-21576) |

